# Cannabis-Microbiome Interactions in Varied Clinical Contexts: A Comprehensive Systematic Review

**DOI:** 10.1101/2022.12.31.22284080

**Authors:** May Soe Thu, Thunnicha Ondee, Szaye Rawicha Hall, Tanawin Nopsopon, Ananya Jagota, Nattiya Hirankarn, Joanne L. Fothergill, Barry J. Campbell, Krit Pongpirul

**Author notes:** **Corresponding Authors:** Krit Pongpirul and Barry J. Campbell.

## Abstract

**Background:** With cannabis legalization spreading to more countries for both medicinal and recreational use, grasping its effects on the human body is vital. The endocannabinoid system, governed by natural and external cannabinoid compounds, significantly impacts host metabolism. Working alongside the host’s immunomodulation, it shapes the gut microbiota, yielding benefits for the GI and immune systems.

**Objective:** To assess the link between cannabis treatment and the gut/oral microbiome.

**Methods:** We extensively searched PubMed, Embase, and Cochrane Library’s CENTRAL until December 9, 2023, for English studies involving adults with clinical abnormalities. Identified studies were analyzed, categorizing by different clinical aspects. Data was then qualitatively and quantitatively synthesized.

**Result:** The study involved 10 studies encompassing 2511 participants, comprising 2 clinical trials and 8 observational studies. The review provided a range of microbiota by the influence of cannabis usage within different clinical contexts: HIV infection, pain/inflammation, systemic aspergillosis, obesity, cognitive deficits, and oral diseases. Users with anhedonia and HIV infection showed lower α-diversity, but those with knee arthritis showed higher α-diversity. According to research, 21.4% of MJ cigarette users experienced adverse outcomes; however, these lessened once they stopped smoking.

**Conclusion:** These findings shed light on the complex effects of cannabis use on the human microbiota, underscoring the need for future research on the therapeutic potential of cannabis. This review provides valuable insights to guide future investigations in this field.

**Registration ID:** PROSPERO 2022 CRD42022354331

**Short Summary:** With the global expansion of cannabis legalization, understanding the effects of cannabis on the human body, particularly among individuals with diverse clinical conditions, is of paramount importance. Through a meticulous systematic review utilizing comprehensive data, our findings uncover that cannabis consumption in adults with varied clinical conditions leads to discernable alterations in the human microbiome. These noteworthy modifications necessitate careful consideration in future investigations exploring the potential beneficial or adverse effects of cannabis treatment on patients.

## Introduction

Marijuana (MJ), or *Cannabis sativa*, has a long history of use both for therapeutic and recreational purposes. It is known as cannabis and comprised of a complex mixture of natural cannabinoids. Cannabidiol (CBD), a non-psychoactive cannabinoid molecule, had its chemical structure first determined in 1963, and the psychoactive δ9-tetrahydrocannabinol (THC) was then discovered in 1964 (1). Cannabis is used in various forms, such as smoking, inhaling, and as cannabis extracts, and ranks as the third most commonly used psychoactive substance worldwide, following alcohol and cigarettes (2). In 2020, the global cannabis user population was over 4%, and nearly 6% among ages (3). Intriguingly, its consumption in population aged ≥ 50 years has been elevated from 15.1% in 2014 to 23.6% in 2016 since legalization for medical use encouraged former non-users to start using it (4). With an expansion of legalized countries, more research studies are being explored on its potential therapeutic effects and adverse outcomes.

The effects of cannabis are mediated via the endocannabinoid system (ECS), which is composed of endogenous ligands such as anandamide (AEA) and 2-arachidonoylglycerol (2-AG), anabolic/catabolic enzymes such as fatty acid amide hydrolase (FAAH, for anandamide) and monoacylglycerol lipase (MAGL, for 2-AG), and endocannabinoid receptors (eg. CB1 and CB2). Although the ECS has been linked to immunological, metabolic, and nervous system homeostasis in addition to playing a regulatory role through the gut-brain axis, the precise physiological function is still being studied (5-8). Nonetheless, numerous active studies on cannabinoids are underway since there is evidence to support their potential efficacy in treating cardiovascular disease, cancer, and inflammation (9). In addition, it has been used for millennia to treat gastrointestinal (GI) tract inflammation and functional problems, such as cramps, nausea, vomiting, diarrhoea, and stomach pain (10, 11).

Even though people who use medical cannabis have more possible advantages than those who don’t, there are still adverse effects (12). The serious adverse events are categorized into respiratory, thoracic, and mediastinal disorders, gastrointestinal (GI) disorders, nervous system disorders, cerebrovascular disorders, general disorders and administration-site conditions, renal and urinary disorders, neoplasm, psychiatric disorders and others. Furthermore, the most frequent physical health reasons are to manage pain (53%), sleep (46%), headaches/migraines (35%), appetite (22%), and nausea/vomiting (21%), while the most prevalent mental health reasons are anxiety (52%), depression (40%), and PTSD/trauma (17%) (13).

The diverse population of the gastrointestinal (GI) microbes engages in a mutually beneficial relationship with the host. These gut microbiota affects neurological, endocrine, and immunological networks through the gut-brain axis and the bilateral communication between the central and enteric nerve systems (14, 15). A mice model shown that the genus *Bifidobacterium* has an antidepressant effect that is partially mediated by microbiome regulation (16). According to a comprehensive analysis of the gut microbiota in anxiety disorders, several taxa and their modes of action may be connected to the pathophysiology of depression and anxiety by communicating with the brain through peripheral inflammation (17). *A. muciniphila* significantly regulates the gut barrier and processes roughly 3–5% of the gut microbiota in healthy humans (18). This demonstrates how closely the GI system and the brain are related.

The endocannabinoidome (eCBome), which encompasses a more extensive network of lipid signaling molecules and receptors related to endocannabinoid function, plays a significant role in the relationship, primarily through CB1 and TRPV1 channels in myenteric and vagal fibres, as well as PPAR-a and GPR119 receptors in enteroendocrine epithelial cells of the small intestine (19). These receptors influence the release of GI neuropeptides, the activity of myenteric neurons, and the function of the vagal and sympathetic nervous systems, all of which may modify the ECS (19). A study highlighted that N-acyl amide synthase genes in gastrointestinal bacteria produce lipids interacting with GPCRs that regulate GI physiology, and these bacterial GPR119 agonists impact metabolic hormones and glucose regulation similarly to human ligands (20). Significant changes in the level and/or composition of the gut microbiota, as observed in germ-free or antibiotic-treated mice, have been demonstrated to impact the messenger RNA (mRNA) expression of eCBome receptors and enzymes as well as the concentrations of eCBome mediators in the gut through still-yet-unknown pathways (21). Additionally, a study discovered that the small intestines of mice given antibiotics had lower levels of N-oleoyl and N-arachidonoyl-serotonin, indicating an interaction that may have an impact on the gut-brain axis.

While examining the effectiveness of THC therapies, *Ruminococcus gnavus*, a good bacterium, was discovered to be more prevalent and pathogenic *A. muciniphila* in the gut and lungs was decreased along with the enrichment of propionic acid (22). Another study found that combining THC and CBD reduced the signs of experimental autoimmune encephalomyelitis (EAE), which was characterized by an increase in anti-inflammatory cytokine production, a decline in pro-inflammatory cytokines, a reduction in mucin-degrading *A. muciniphila*, and a reversal of the high level of lipid polysaccharides (23). Collectively, these findings indicate that cannabis affects the gut microbiome.

The current body of literature lacks a comprehensive and systematic synthesis of research on the intricate relationship between cannabis and the microbiome across diverse clinical contexts. Existing studies have explored aspects of this interaction, but there is a notable absence of a unified and rigorous analysis that integrates findings from different clinical scenarios. A systematic review can identify gaps in the coverage of clinical conditions and highlight areas where more research is needed to understand the cannabis-microbiome interaction comprehensively. To promote more in-depth research, this review will be the first to assess the microbiome of multiple observational studies and clinical trials including cannabis/MJ use in humans with a variety of disorders.

## Materials and Methods

### Protocol and registration

The systematic review was registered on PROSPERO ID 2022 CRD42022354331.

### Literature search, study selection and data extraction

In compliance with the PRISMA declaration standards, the review was conducted (24). The studies, which were published up until December 9, 2023, were imported from the databases of PubMed, Embase, and CENTRAL. The search was specifically limited to studies that addressed microbiomes and cannabinoids, as well as the impacts of cannabis treatment and how it affects the microbiome.

Interventions examining the effects of any cannabis treatment and its influence on microbiome change, with or without active or placebo controls, are reflected in the inclusion criteria. Animal studies, in vitro research, procedures, reviews, opinions, letters, comments, and guidelines were not included. Data extraction was done after two reviewers independently determined whether the papers met the eligibility requirements based on the abstract and full text screening. Consensus was used by authors to settle any disputes.

The following is how the data extraction procedure was carried out: 1) research attributes include the names of the authors, the year of publication, the type of study, the nation, the sample size, and the age range; The data extraction process involved a thorough examination of pertinent text, tables, and figures. 2) Baseline characteristics, including participant information, study region, and clinical conditions reported by both patients and controls; 3) subgroup evidence, such as microbial profile and diversity, classified by specific diagnostic health problems; and 4) adverse reactions associated with MJ consumption.

### Risk of bias

Using the ROB2 method, the risk of bias (ROB) in the extracted intervention study (25) was assessed. The non-randomized clinical trial (26) utilised the ROBINS-I (Risk Of Bias In Non-randomized Studies -of Interventions), and the cohort and case-control studies (27-34) employed the Newcastle-Ottawa Quality Assessment Scale (NOS).

### Statistical analysis

The prevalence of the characteristics was described by total number and percentage. The overall mean age of included studies was calculated using combined mean and standard deviation (SD) techniques.

## Results

### Study selection

In the initial literature search, 5,000 articles were identified across various databases. Following the removal of 1,533 duplicates and the analysis of the titles and abstracts of the remaining 3,467 studies, 3,428 publications were excluded based on predetermined criteria, leaving 39 articles to undergo full-text screening. Ultimately, the systematic review included 10 studies that met the eligibility criteria (Figure 1).

**Figure 1:**
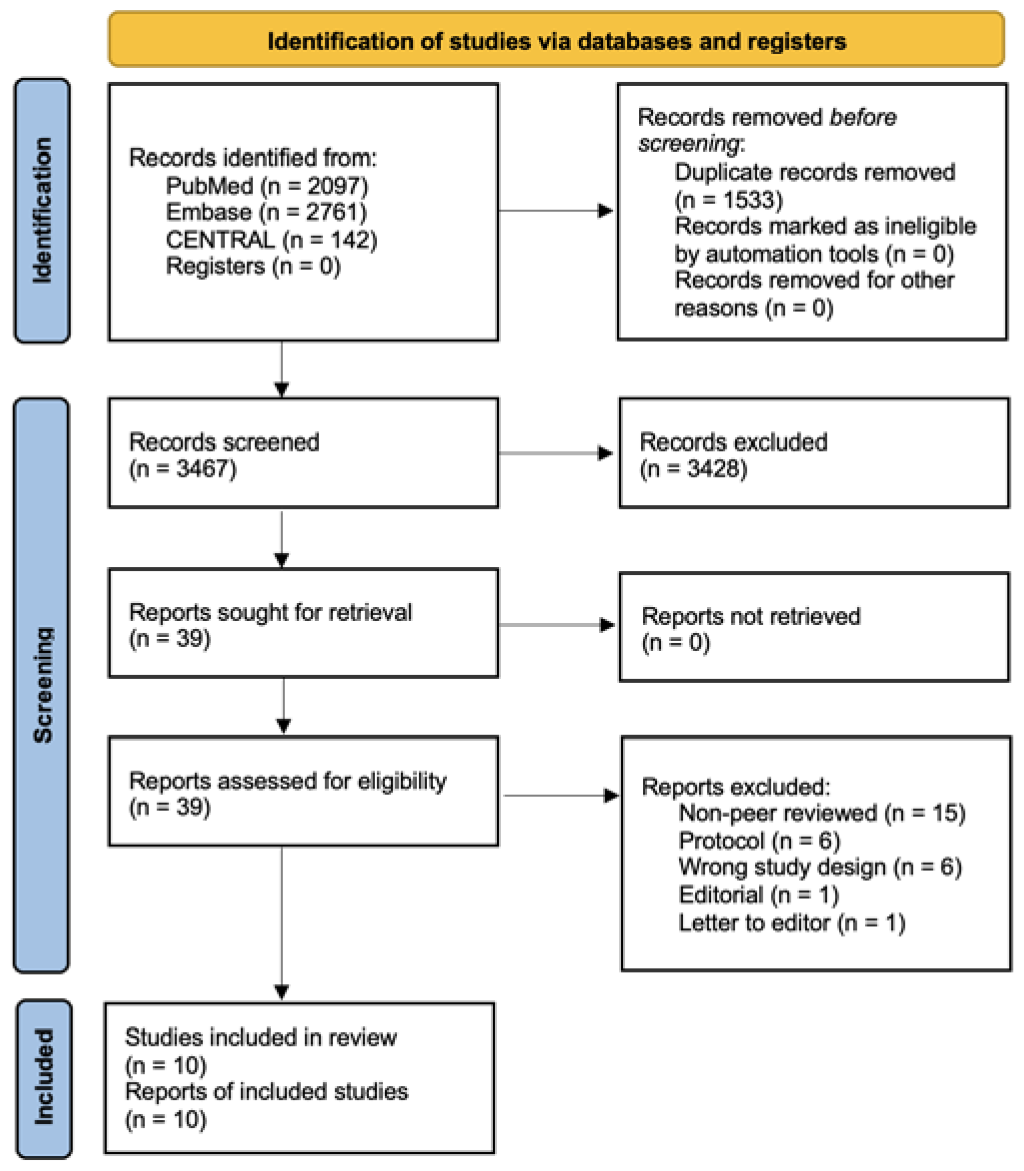
Flow diagram for identifying studies in the systematic literature review.

**Figure 2.**
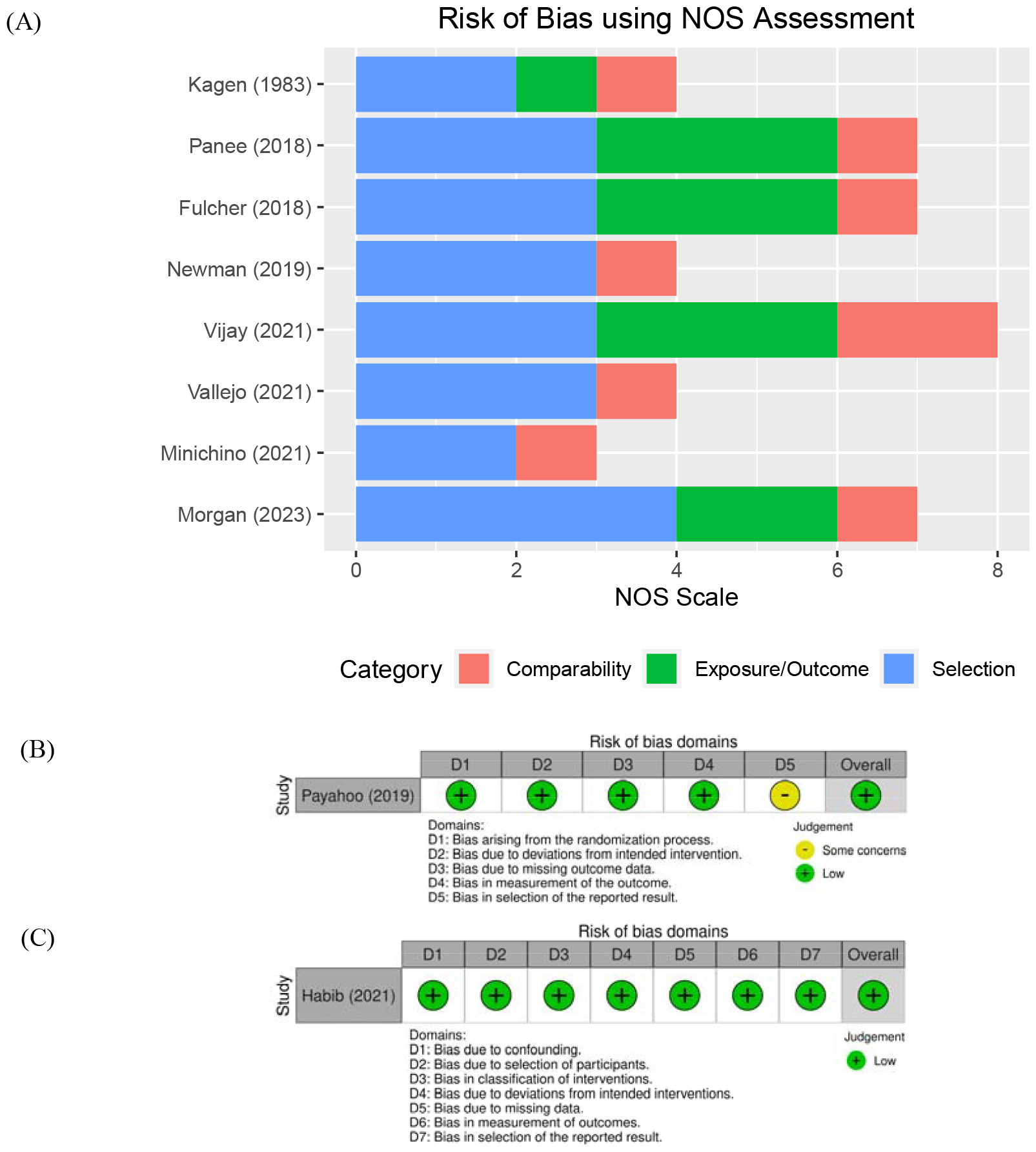
Assessment of risk of bias. **2A**, Using NOS for cohort and case-control studies; **2B**, Using RoB2 for randomized controlled trials; **2C**, Using ROBINS-I for non-randomized studies of interventions. Note: The Newcastle-Ottawa Scale (NOS), version 2 of the Cochrane risk-of-bias tool for randomized trials (RoB 2), Risk Of Bias In Non-randomized Studies of Interventions (ROBINS-I)

### Study characteristics

The included studies were one case-control (31), seven cohort (27-30, 32-34), and two clinical trials (25, 26), at which it involved 2,511 participants ranging from 1983 to 2023 (Table 1). The studies were conducted at four different countries: the United States of America (USA), the Islamic Republic of Iran (Iran), the State of Israel, and the United Kingdom (UK).

**Table 1.** Baseline characteristics of the included studies.

| # | Author | Published year | Study period | Study type | Country | Sample size | Age range | Ref. |
| --- | --- | --- | --- | --- | --- | --- | --- | --- |
| 1 | Payahoo | 2019 | 2016 - 2018 | Randomized double-blind clinical trial | Iran | 56 | 18 - 59 | (25) |
| 2 | Habib | 2021 | 2019 - 2020 | Non-randomized clinical trial | Israel | 16 | 27-78 | (26) |
| 3 | Kagen | 1983 | 1982 | Case-control study | USA | 38 | 17 - 36 | (31) |
| 4 | Panee | 2018 | - | Cohort study | USA | 39 | 21 - 36 | (28) |
| 5 | Fulcher | 2018 | 2014 - 2016 | Cohort study | USA | 37 | 28-39 | (29) |
| 6 | Newman | 2019 | - | Cohort study | USA | 39 | 18 - 58 | (33) |
| 7 | Vijay | 2021 | 2018 - 2020 | Cohort study | UK | 78 | >45 | (27) |
| 8 | Minichino | 2021 | - | Cohort study | UK | 786 | 18 -101 | (30) |
| 9 | Vallejo | 2021 | 2019 | Cohort study | USA | 1380 | 20-34 | (32) |
| 10 | Morgan | 2023 | - | Cohort study | USA | 42 | 16-20 | (34) |

### Subject characteristics

The table 2 outlines the demographic and clinical characteristics of participants in a study, distinguishing between patients and controls. Notably, the patient group, constituting 43% of the total participants, exhibits a broader age range and a higher mean age (57.6 years) compared to the control group, which skews younger with a mean age of 28.4 years. Furthermore, there is a distinct gender imbalance, with a higher percentage of females in both groups. Geographically, the majority of patients are from the UK (76.9%), while controls predominantly hail from the USA (95.2%). Clinically, patients present with diverse conditions, with cognitive deficits being notably prevalent (72.7%).

**Table 2.** Baseline characteristics of the included clinical trials.

| Characteristics | Patients | Controls |
| --- | --- | --- |
| Participants, n (%) | 1071 (43) | 1440 (57) |
| Age range | 16-101 | 16-87 |
| Age, mean (SD) | 57.6 (14.6) | 28.4 (11.3) |
| Gender, n (%) |  |  |
| Male | 198 (17.9) | 79 (5.4) |
| Female | 910 (82.1) | 1371 (94.6) |
| Region, n (%) |  |  |
| USA | 204 (19.0) | 1371 (95.2) |
| Iran | 27 (2.5) | 29 (2.0) |
| Israel | 16 (1.5) | 0 (0) |
| UK | 824 (76.9) | 40 (2.8) |
| Clinical consideration, n (%) |  |  |
| HIV infection | 137 (12.4) | - |
| Pain/Inflammation | 54 (4.9) | - |
| Systemic aspergillosis | 28 (2.5) | - |
| Obesity | 64 (5.8) | - |
| Cognitive deficits | 805 (72.7) | - |
| Oral disease | 20 (1.8) | - |

### Analysis for Risk of bias

Among 7 observational studies (27-33), the study by Minichino et al. displayed a high risk due to the absence of a description of the non-exposed cohort, the inability to blind the outcome assessors, and a lack of follow-up information (Figure 2A). Similarly, previous cohort studies by Newman et al., and Vallejo et al. also exhibited biases due to challenges in outcome assessment blinding and the absence of a follow-up timeline description. In the study by Kagen et al., a significant bias was identified due to a lack of statements regarding case and control selection and exposure. Furthermore, in the reported results of the RCT conducted by Payahoo et al., there was evidence of selection bias as they did not include the specified lipid profile analysis mentioned in their protocol (Figure 2B) while the non-RCT study by Habib et al. demonstrated a well-performed risk assessment (Figure 2C).

### Microbiota alteration by different cannabis usage

After consuming MJ/medical cannabis or its constituents, patients with a range of clinical diseases had their microbiological changes evaluated (Figure 3), and the qualitative synthesis of the microbial diversity and related parameters was conducted (Table 3). Three categories of cannabis use were identified in this review: THC, endocannabinoids, and MJ.

**Table 3.**
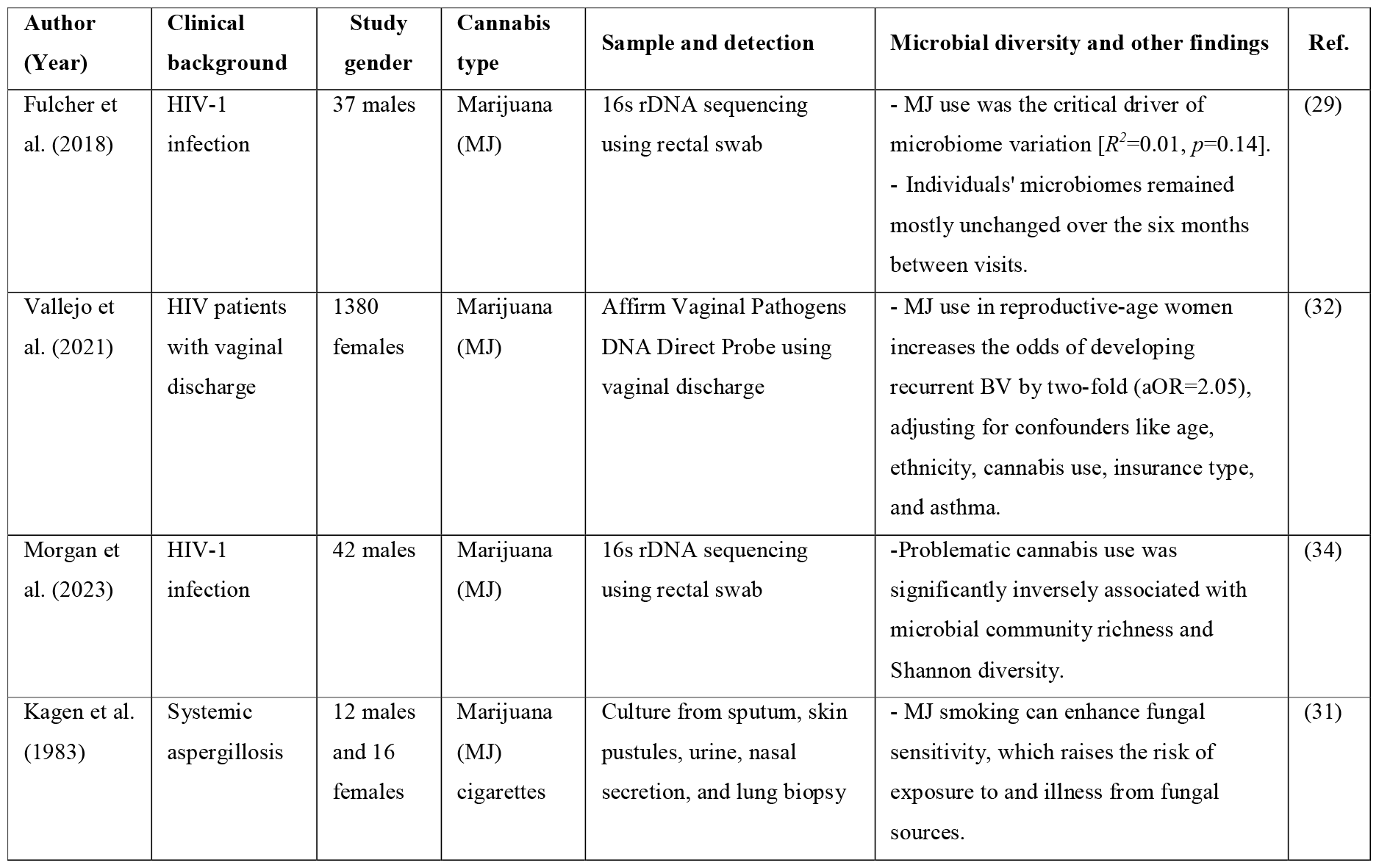
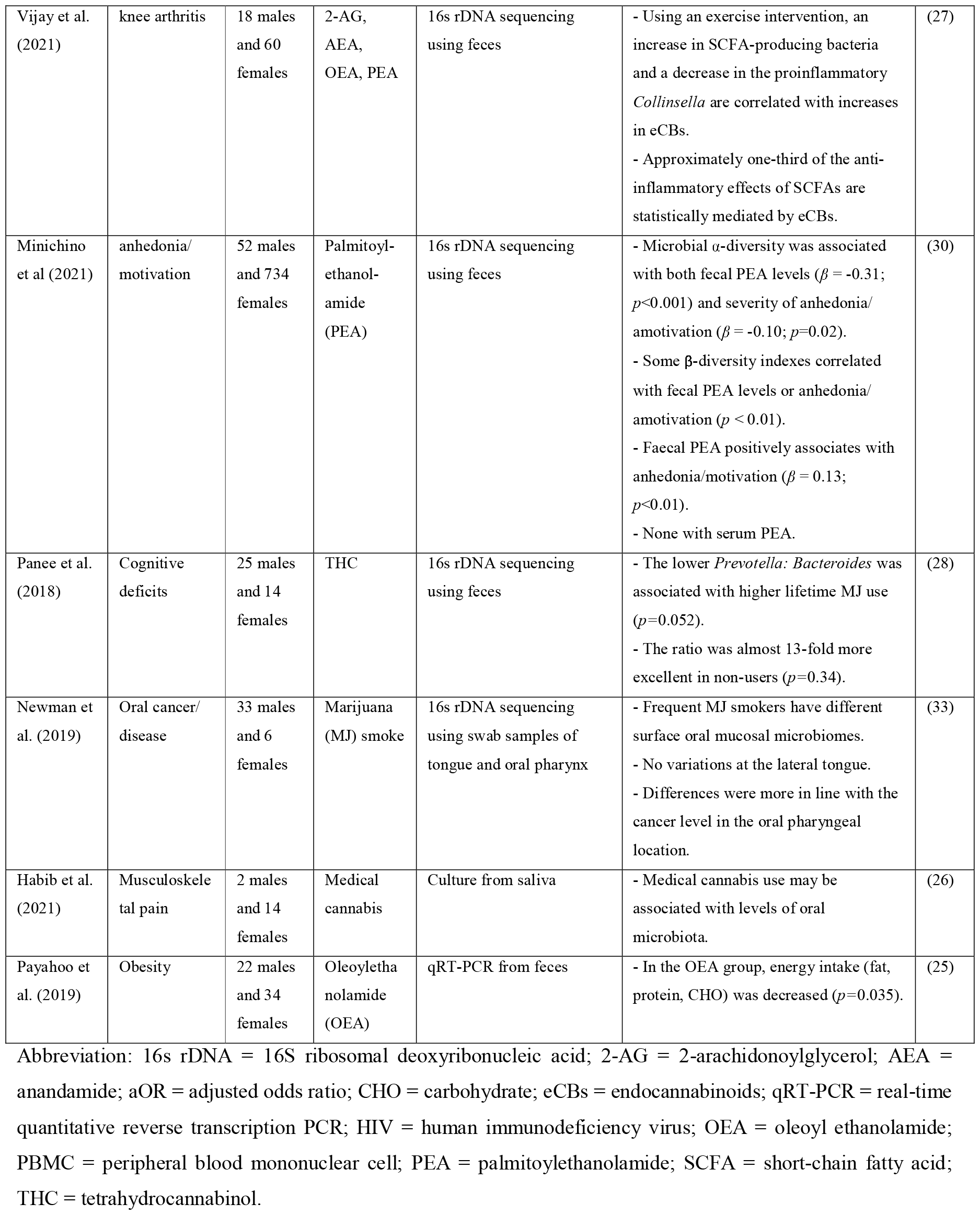
Qualitative analysis of microbial changes on the use of cannabis.

**Figure 3.**
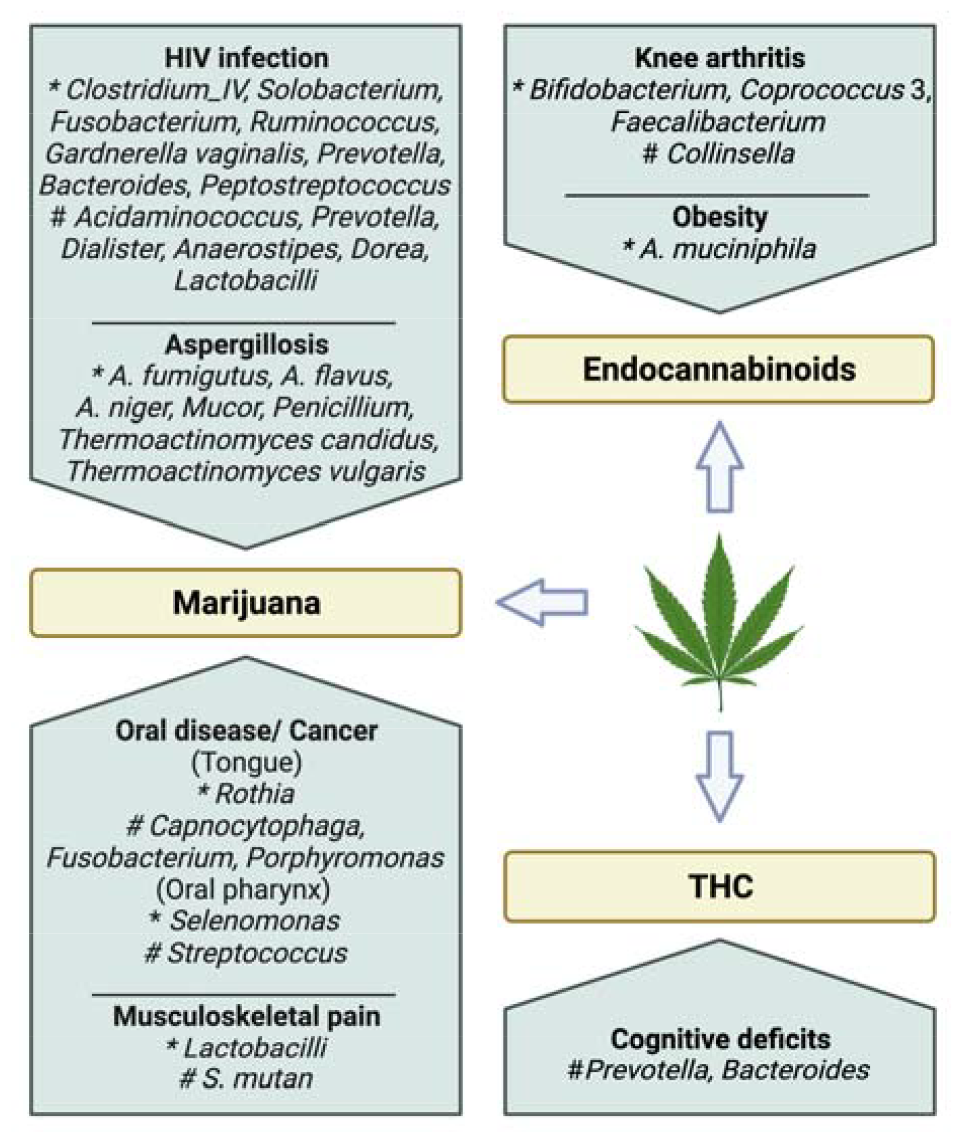
Positive and negative associations of microbiome profiles in varied clinical conditions, examining the impact of different forms of cannabis use *: positive association; #: negative association; THC: tetrahydrocannabinol.

### Adverse events

Six patients (21.4%) reported symptoms, including coughing, and wheezing after using MJ cigarettes, while one (3.5%) suffered drowsiness, night sweats, systemic aspergillosis, and coughing bouts. Steven et al. reported these negative consequences, which went away quickly after quitting smoking MJ (31).

## Discussion

In this systematic review, the study encompassed patients with oral disease, obesity, systemic aspergillosis, pain/inflammation, and HIV infection, employing MJ through substance use, oral capsules, or cigarette smoking. There exists an ongoing debate regarding the efficacy of probiotic treatments in altering microbiota composition (35). The limitations of the 16S rRNA sequencing method, often considered less sensitive to minor microbiota alterations, contribute to this uncertainty. However, it is crucial to emphasize that significant changes in gut microbiota composition may not be an exclusive prerequisite for substances to confer health benefits. Rather, health benefits can manifest through the production of metabolites and intricate interactions with the host’s metabolism and immune system, even in the context of various disease states. In light of these considerations, a comprehensive systematic review on the interaction between cannabis and the microbiome is imperative to provide a nuanced understanding of the potential health implications stemming from this intricate relationship.

### Microbial alteration in HIV patients using MJ

A study by Fulcher et al., which focused on HIV-positive men who have sex with men (HIV+ MSM) (29), showed that the patients’ rectal swab samples had a high relative abundance of *Prevotella*. Additionally, there was a positive correlation between MJ use in HIV patients and elevated levels of *Fusobacterium* and *Anaerotruncus*, coupled with a negative association with *Dorea*, as describe in Figure 3. Evidence has shown that intestinal dysbiosis, marked by a decrease in *Bacteroides* and an increase in *Prevotella*, is linked to HIV infection (36, 37). According to recent research, elevated *Prevotella* in HIV may be a contributing factor to the gut’s ongoing inflammation, which can cause mucosal dysfunction and systemic inflammation (38, 39). Marijuana usage in HIV+ MSM was linked to lower *Prevotella* abundance when propensity score analysis was used to account for several covariates (29).

There is evidence that HIV patients also had decreased levels of *Lactobacillus* and other beneficial microbes, as well as an increase of potentially opportunistic infections (37). Additionally, MJ users among HIV patients have been verified by Vallejo et al. to engage in higher-risk sexual behaviour, which can result in bacterial vaginosis (BV), which is characterised by an overgrowth of facultative anaerobic organisms and the absence of *Lactobacilli* (32). The prevalence of trichomonas infection in rectal samples decreased from 14% to 5% over a half-year (*p*=0.08), while gonorrhea and syphilis infections increased from 8% to 11% (*p*=0.66) and 0% to 5% (*p*=0.16), respectively (29). Intriguingly, MJ users were over six times more likely to test positive for *T. vaginalis* (aOR=6.2, *p*=0.0003), indicating a potential association with sexually transmitted diseases (40). The analysis demonstrating a significant association between MJ use and recurrent BV also presented an adjusted odds ratio of 2.05 (32), as outlined in Table 3. Despite these findings, additional clinical studies are warranted to address the ongoing controversy surrounding the relationship between MJ use and changes in sexually transmitted infections.

Interestingly, Fulcher et al. also noted that 28.4% of MJ users had a history of asthma, contrasting with 18.3% of non-users (*p*<0.01) (29). Despite the bronchodilator effect of cannabis on the airway, suggesting potential benefits for asthma patients, there are acknowledged detrimental effects on the lungs (41). This dual impact prompts consideration of cannabis use for medicinal or recreational purposes, especially considering reported improvements in asthma symptoms.

Additionally, a study group (34) found a substantial adverse relationship between shannon diversity and microbial community richness and problematic cannabis usage. These findings collectively highlight the intricate associations between cannabis use, microbiota alterations, and various health outcomes in HIV patients, emphasizing the need for further research to elucidate the underlying mechanisms and potential therapeutic implications.

### Impact of MJ on microbiota regarding pain or inflammation

The ability of AEA and THC to raise the levels of AMPs and SCFAs in mouse models of inflammation has been confirmed by numerous studies (42, 43). The main SCFAs implicated in host-bacterial contacts are butyrate, generated by Firmicutes, and acetate and propionate, produced by Bacteroidetes (44). It’s interesting to recall that dendritic cells and adipose tissue macrophages express GPCRs like GPR109A and GPR43. Colon cancer cells undergo apoptosis when GPR109A is activated in a butyrate-dependent manner (45). Moreover, these receptor/ligand complexes prevent the activation of nuclear factor-kappaB in mice’s colons. HDAC inhibition produced by butyrate inhibited the production of pro-inflammatory cytokines like IL-6 and IL-12 that are stimulated by lipopolysaccharides (46). Numerous studies have demonstrated that SCFAs are the primary mediator between the gut microbiome and host immunological homeostasis. A study conducted by Vijay et al. (27) revealed a positive association between changes in AEA and butyrate while elevations in AEA and palmitoylethanolamide (PEA) were correlated with a decline in TNF-_α_ and IL-6. They also revealed that AEA and OEA at baseline were associated with a higher α-diversity (p = 0.002 and p <0.001, respectively). This suggests the involvement of the ECS in the anti-inflammatory actions of SCFAs, indicating the potential role of additional pathways in the regulation of the immune system by the gut microbiota.

Musculoskeletal pain, a prevalent cause of chronic non-cancer pain, has led patients to perceive cannabis as beneficial for pain relief with minor adverse effects and an improvement in psychological well-being (47). In another trial utilizing medical cannabis, notable changes were observed in the microbial composition. Specifically, there was an elevation of *S. mutans* and *Lactobacilli* in the fourth week, despite being low in the initial week (26). The overall impact of cannabis on these bacteria appeared to exhibit both favorable and unfavorable effects. These findings underscore the intricate relationship between cannabis use, microbial alterations, and the complex interplay with pain perception.

### Different expression of microbiota in oral diseases after the use of MJ

Newman et al. found that genera earlier shown to be enriched on head and neck squamous cell carcinoma (HNSCC) mucosa, such as *Capnocytophaga, Fusobacterium*, and *Porphyromonas*, were at low levels at the tongue site in MJ users, while *Rothia*, which is found at depressed levels on HNSCC mucosa, was high (33). At the oral pharynx site, differences in bacteria were distinct, with higher levels of *Selenomonas* and lower levels of *Streptococcus*, as seen in HNSCC. In samples taken from the lateral border of the tongue and the oral pharynx, it indicated that daily/almost daily inhalation of MJ over the previous month correlates with differently abundant taxa of the oral microbiome. Even though the use of MJ is associated with microbial changes, it is hard to conclude that it is the cause of how normal tissue develops into disease and then SCC. Furthermore, these changes were not consistent with malignancy.

### Gut microbiota in cognitive deficits with the use of MJ

Moreover, using MJ is associated with alterations in gut microbiota and mitochondrial function, leading to cognitive deficits (28), as well as lower fruit and vegetable intake and greater animal-based food consumption. It also found that a more extensive lifetime MJ use was associated with a lower *Prevotella: Bacteriodes* ratio, as indicated in Table 3. Overall, the authors suggested that MJ use and associated dietary change like low dietary intake of antioxidants and fibers, contribute to the dysbiosis.

### Effect of cannabinoids in obesity in terms of microbiome

Obesity is an excessive buildup of fat and cause an inflammatory response. Currently, herbal remedies like *Cannabis sativa* derivatives are gaining popularity in the treatment of obesity and its co-morbidities. The trial discovered that OEA supplement use significantly decreased the energy, fat, protein and carbohydrate intake of obese participants (*p*<0.001) (25). *A. muciniphila* increased considerably compared to the placebo group, suggesting that OEA could be used as a supplement for obese people (47).

### Dysbiosis in cognitive deficits using MJ

Panee et al. (28) revealed that mitochondrial function correlated positively with Fluid Cognition and Flanker Inhibitory Control and Attention scores in MJ users but not in non-users (interaction *p*=0.0018–0.08). So, both mt dysfunction and gut dysbiosis also affect cognition.

## Conclusion

In conclusion, the systematic review on the cannabis-microbiome interaction in varied clinical contexts is crucial for consolidating existing knowledge and identifying key research gaps. By critically analyzing the available literature, this review aims to provide a comprehensive understanding of how cannabis influences the microbiome across different clinical scenarios. Moreover, the identification of methodological variations in current studies will allow for recommendations on standardized approaches. In contrast, it is critical to recognise the several flaws in the review. To begin with, there were just two intervention studies at which one was non-randomized. As a result, the study was limited to assessing the impact of cannabis therapy. Second, this was merely a data presentation in the current study because the adverse event data were only discovered through a study. Finally, while the study addressed a variety of clinical circumstances, only the qualitative evaluation could be completed. Despite the limited literature dedicated to these interactions, this should be anticipated for further work in this exciting research field. The systematic review will serve as a valuable resource for researchers, healthcare professionals, and policymakers, offering insights into the potential therapeutic implications of the cannabis-microbiome interaction and informing the development of targeted interventions across a spectrum of clinical conditions.

## Data Availability

All data produced in the present work are contained in the manuscript as supplementary files.

## Abbreviations

CENTRAL: Cochrane Central Register of Controlled Trials
CIs: Confidence intervals
GRADE: Grading of Recommendation Assessment, Development and Evaluation
NOS: Newcastle - Ottawa Quality Assessment Scale
PRISMA-P: Preferred Reporting Items for Systematic review and Meta-analysis Protocols
RCTs: Randomized controlled trials
ROBINS-I: Risk Of Bias In Non-randomized Studies - of Interventions
US: United States.

## Acknowledgements

None

## Authors’ contributions

MT and SR contributed to the analysis and writing of the manuscript. MT, TO, TN, SR and KP contributed to the conception and design. MT, SR, and BC conducted the data curation. MT, BC, JF, NH, and KP contributed to the critical revision of the manuscript. All authors read and approved the final manuscript. KP is the guarantor of the review.

## Availability of data and materials

Data will be available as supplementary files.

## Ethics approval and consent to participate

Not applicable.

## Consent for publication

Not applicable.

## Patient and Public Involvement

Patients or the public were not involved in the design, or conduct, or reporting, or dissemination plans of our research.

